# Comparative ultrasonographic anatomy of “medial-oblique” probe position with classic “short axis” probe position for ultrasound guided axillary vein cannulation: A cross-over study

**DOI:** 10.1101/2022.06.01.22275893

**Authors:** Souvik Maitra, Sulagna Bhattacharjee, Dalim K. Baidya, Bikash R Ray

## Abstract

**Study objective:** Subclavian vein cannulation in the infraclavicular region may be a preferred approach as it is associated with less infection than internal jugular or femoral venous access in critically ill patients. ‘Short-axis view” has a problem that shaft of the puncture needle is not visualized and tip of the needle is only seen as ‘dot’; hence posterior wall of the vein puncture is always possible with ‘short-axis approach’. We have designed this study to know the ultrasonographic anatomy of subclavian vein in short axis and medial oblique view.

**Design:** Prospective randomized, cross-over.

**Setting:** Operating room.

**Patients:** One hundred consecutive patients aged between 18-50yrs of either sex, ASA PS I-III undergoing elective surgery under general anesthesia.

**Intervention:** The ultrasonographic anatomy of axillary vein cannulation in infraclavicular region in one hundred patients was assessed as per randomization sequence in medial oblique probe position and short-axis probe position.

**Measurements:** Antero-posterior and transverse diameter of axillary vein, pleural line visibility.

**Main Results:** Transverse diameter of subclavian vein was higher in ‘medial-oblique’ view [mean difference (95% confidence interval) 0.12 cm (0.09-0.16) cm; p<0.0001]. However, mean (standard deviation) antero-posterior diameter was similar in two views [0.72(0.15) cm in short axis versus 0.68 (0.16) cm in medial oblique; p=0.07]. Visibility of pleural line was similar in both the view (p=0.84, Chi-square test).

**Conclusion:** Medial-oblique view may be useful in axillary approach of subclavian vein cannulation. Further clinical studies are required to know whether medial oblique view can reduce complications and increase success rate of ultrasound guided subclavian vein cannulation over short axis view.

---

Central venous cannulation is one of the commonest procedures performed in the intensive care unit as well as in the operating room. Subclavian vein cannulation in the infraclavicular region may be a preferred approach as it may be associated with less infection than internal jugular or femoral venous access in critically ill patients^1^. Subclavian venous access is also required in the operating room in patients undergoing head and neck surgeries. Classic ultrasound guided approach of Subclavian vein cannulation is basically a trans-pectoral axillary vein cannulation^2^ and most important complication associated with subclavian vein cannulation is pneumothorax and arterial puncture^3^. Both short axis^4^ and long axis approach^5^ for ultrasound guided subclavian vein cannulation have been described in the literature. The ‘short-axis view” has an inherent problem that shaft of the puncture needle is not visualized and tip of the needle is only seen as ‘dot’; hence posterior wall of the vein puncture is always possible with ‘short-axis approach’. On the contrary, in long axis view, vein and artery are usually overlapping and pleural margin is often not seen. DiLisio et al^6^ in 2012 described a new “medial oblique technique” for ultrasound guided internal jugular vein (IJV) cannulation. The authors reported that this approach allows for optimal imaging of the IJV and the carotid artery side by side and following the needle throughout the insertion from skin to vessel penetration in a medial-cephalad to lateral-caudad direction. This technique combines the advantages of the short-axis and long-axis approaches and minimizes the risk of carotid puncture from a medial-to-lateral needle direction. Subsequently, Baidya et al^7^ compared sono-anatomy of medial oblique view and short axis view for internal jugular vein cannulation. In this study we assessed anatomical feasibility of medial oblique approach and compare with ‘short axis’ approach for infraclavicular subclavian vein cannulation.

## Methodology

After obtaining written informed consent & permission from the institute ethics committee, one hundred consecutive patients aged between 18-50yrs of either sex and ASA PS I-III undergoing elective surgery under general anesthesia was recruited in this randomized cross over study. This study was registered in the National Clinical Trial Registry of India (Registration no: CTRI/2017/12/010765; www.ctri.nic.in).

Following patients were excluded from this study:

I. Refusal for consent to participate
II. ASA PS> III
III. Pregnancy
IV. BMI > 30.00 or BMI< 18.00
V. Patients with distorted neck anatomy and previous neck surgery or chest wall surgery.

### Randomization & blinding

A computer-generated random number list was used to prepare serially numbered opaque envelopes that will contain the details of one of the probe position to be followed first. The sealed envelope was handed over to the anaesthetic team (not part of the investigating team) who then opened and followed the mentioned technique. An anaesthesiologist who is unaware about the group to which the pictures belong will analyze data obtained from the ultrasound-generated image. The sono-anatomy of axillary vein cannulation in infraclavicular region in one hundred patients will be assessed as per randomization sequence in medial oblique probe position and short-axis probe position.

Group L= Sono-anatomy of axillary vein in infraclavicular region in “medial oblique approach” probe position

Group S= Sono-anatomy of axillary vein in infraclavicular region in “short-axis” probe approach

### Sample size

No previous study has evaluated either of the probe position in terms of anatomical relationship. We analyzed 100 views in each group to obtain normally distributed data.

### Technique

After approval of institute’s ethical committee and obtaining written, informed consent from the patients, 100 patients will be enrolled for this study. Patients will undergo thorough preoperative evaluation and will be checked against the exclusion criteria of this study. In the operating room, after induction of general anesthesia, patients infraclavicular area was draped in the usual sterile fashion and placed in the 30-degree trendelenburg position with the patient’s head turned 15 degrees to the left side. A linear ultrasound probe was covered in a sterile sleeve and placed below the clavicle at the junction of medial two third and lateral one third of clavicle. Once when the short-axis view was obtained, and required measurements were noted. Then the probe was rotated approximately 30° anti-clockwise, in a medial-cephalad to lateral-caudad direction. This positioning will allow for visualization of the axillary vein in more of a long-axis view and imaging of the axillary artery at the medial aspect of the image displayed in its short axis. Then again ultrasound image will be frozen and required analysis will be done.

Either “short-axis” view or “medial oblique” view was obtained first in each patient, as per sequence generated by a random number table. Data from the images obtained in “short-axis” view were analyzed as Group S (n=100) and those images were obtained in “medial-oblique” view were analyzed as Group L (n=100).

Following data were collected from each group of images:

I. Relative position of axillary vein in relation to axillary artery (medial, posterior or postero-medial)
II. Length of overlapping of axillary vein over axillary artery
III. III. Transverse and antero-posterior diameter of axillary vein

### Plan for analysis of data

Demographic data were expressed as mean ± SD (age, weight, height) or proportion (sex and ASA physical status). Relative position of axillary vein with axillary artery was expressed as proportion. Continuous variables were analyzed to two-tailed independent sample t-test. Qualitative data were compared using Chi-square test. A p-value of less than 0.05 was considered as statistically significant.

## Results

Images of subclavian vein were obtained in all 100 patients both in short axis and medial oblique view. Mean (SD) age of the recruited patients were 49.4 (9.6) y, mean (SD) body weight was 55.8 (7.2) kg and 54% patients were male. Transverse diameter of subclavian vein was significantly higher in ‘medial-oblique’ view [mean (SD) 0.69 (0.11) cm in short axis view versus 0.81 (0.14) cm in medial oblique view; mean difference (95% confidence interval) 0.12 cm (0.09-0.16) cm; p<0.0001]. However, mean (standard deviation) antero-posterior diameter was similar in two views [0.72(0.15) cm in short axis versus 0.68 (0.16) cm in medial oblique; p=0.07]. Visibility of pleural line was similar in both the view (p=0.84, Chi-square test).

## Discussion

Most important finding of our study is that both the subclavian vein and artery were visible in oblique axis view and transverse diameter of the SCV was higher in the oblique axis view. Incidence of overlapping was also less in oblique axis view.

With best of our knowledge, no previous study has evaluated oblique axis view of infraclavicular SCV cannulation. De Cassai & Galligioni reported that they use oblique approach for SCV cannulation regularly in their practice; but they did not report any data in that article^8^. Baidya et al. reported feasibility of medial oblique approach for IJV cannulation^7,9^. Results of our study is similar to the previous study by Baidya et al., who reported that transverse diameter of IJV is higher in medial oblique position with a reduction in overlap between IJV and carotid artery^7^.

Short-axis view allows the visualization of both SCV and SCA with the echogenic pleural line posterior and inferior to the artery and vein. However, short axis view does not allow the operator to visualize the whole length of needle being inserted; it only allows to visualize the needle tip as a ‘dot’^9^. On the other hand, though long axis view allows to visualize the SCV as a hypoechoic longitudinal structure; but it is not possible to identify SCA in long axis view as a separate structure. Oblique axis view offers advantages of both long axis and short axis approach. In medial oblique view, the ultrasound beam is transmitted through an oblique axis over the SCV. So, a longer length of SCV is being scanned in oblique axis, which increased effective transverse diameter. Baidya DK et al. reported that oblique axis approach improved needle tip and shaft visibility during IJV cannulation^9^. Pleural line visibility, which is one of the most important landmarks for safe SCV cannulation, was present both in short axis and medial oblique view.

Complications after subclavian vein cannulation is around 9.7% and a large randomized controlled trial reported that ultrasound guided localization of SCV before cannulation does not reduce incidence of procedure related complications^10^. However, real time USG guidance in longitudinal for SCV cannulation offers a higher success rate and lower rate of complications including less arterial puncture and pneumothorax. However, despite of USG guidance, catheter misplacement was similar.^5^ We believe oblique axis view USG guided SCV cannulation needs further clinical evaluation.

## Limitations

As our study only evaluated anatomical characteristics of SCV and SCA, it is not possible to delineate whether medial oblique approach offers advantages over short axis approach for SCV cannulation

## Conclusion

Medial oblique view for infraclavicular SCV cannulation may offer anatomical superiority over short-axis approach as transverse diameter is increased and overlapping is reduced in the former view. Further clinical studies are required to identify whether medial oblique approach is actually a safer alternative to short axis approach or not.

## Data Availability

All data produced in the present study are available upon reasonable request to the authors

---

**Figure 1:**
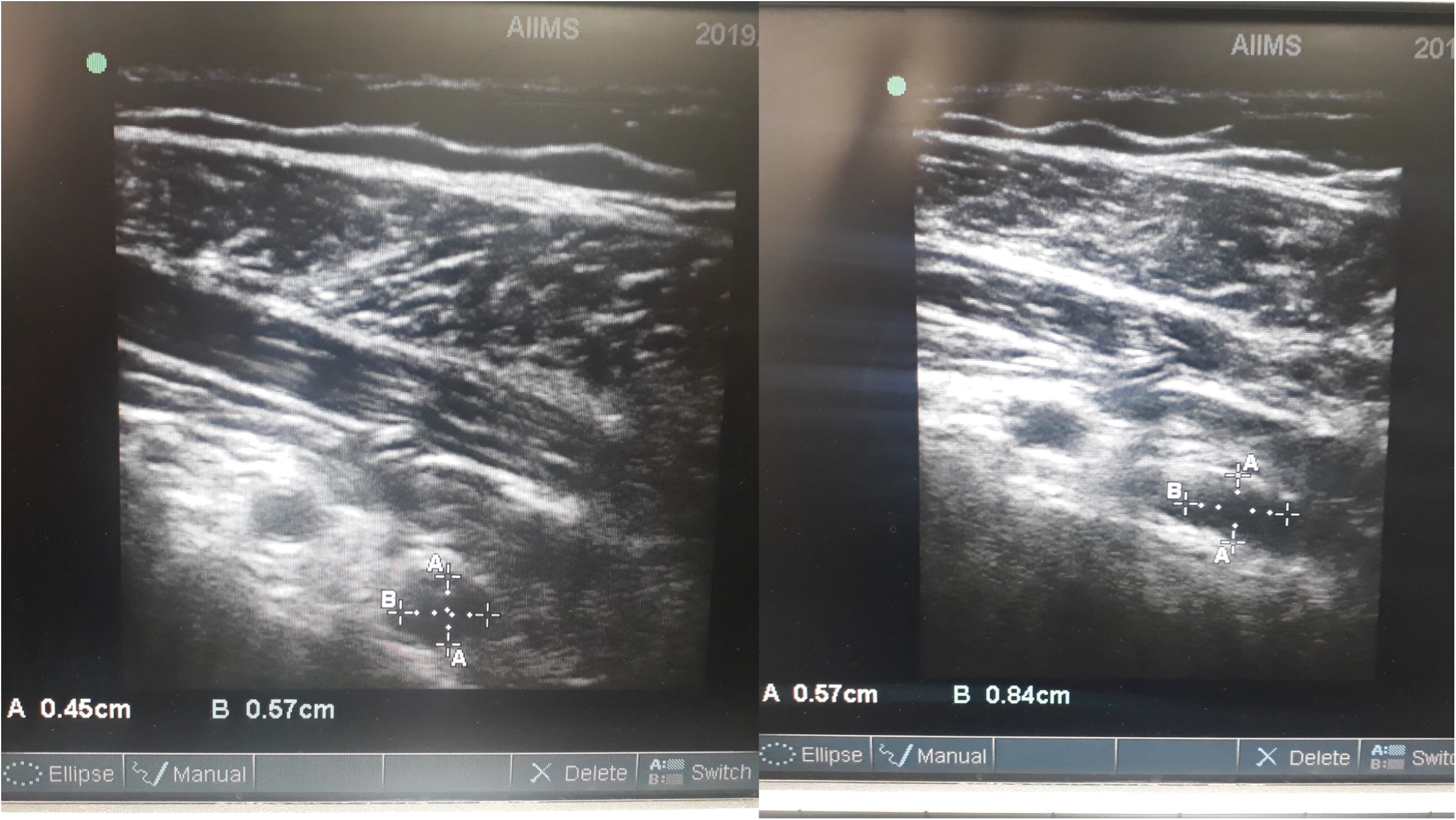
Measurements of antero-posterior and transverse diameter of axillary vein in short axis (left) and medial oblique view (right).

## Notes

### Competing Interest Statement

The authors have declared no competing interest.

### Clinical Trial

CTRI/2017/12/010765

### Funding Statement

No funding was received

### Author Declarations

This study was approved by the Institute Ethics Committee, All India Institute of Medical Sciences, New Delhi, India (vide permission letter no: IEC- 580/05.01.2017).

